# Inclusion, characteristics and credibility of systematic reviews in doctoral theses: A cross-sectional study of all Medical Faculties in Sweden

**DOI:** 10.1101/2024.06.11.24307851

**Authors:** M. Ringsten, K. Färnqvist, M. Bruschettini, M Johansson

**Affiliations:** Cochrane Sweden, Lund University, Skåne University Hosptial, Skåne, Sweden; Molecular Medicine and Surgery, Karolinska Institute, Stockholm, Sweden; Paediatrics, Department of Clinical Sciences Lund, Lund University, Skåne University Hospital, Lund, Sweden; Global Center for Sustainable Healthcare, School of Public Health and Community Medicine, Sahlgrenska Academy, University of Gothenburg, Gothenburg, Sweden

**Keywords:** Systematic review, scoping review, doctoral thesis, PhD thesis, doctoral education, AMSTAR-2, risk of bias

## Abstract

**Objective:** Systematic reviews (SRs) are essential to ensure that decisions are informed by an up-to-date and complete understanding of the relevant research evidence. Conducting SRs within a doctoral thesis can reduce redundant, harmful and unethical research, identify knowledge gaps, and help the doctoral student obtain important skills to conduct and use research. The output and learning process of SRs overlaps with the aims of doctoral programs. We aim to explore to what extent SRs are included in doctoral theses from all medical faculties in Sweden, and to describe the type, topic and assess the credibility of the reviews.

**Study design and setting:** Duplicate assessors independently searched local and national repositories for doctoral theses published in 2021 within all seven medical faculties in Sweden, and categorized identified reviews based on review type, topic, and credibility using AMSTAR-2.

**Results:** 5.4% (45/852) of all doctoral theses included a review, and 1.3% (45/3461) of all included studies were reviews. Of these, two thirds (31) were SRs and the rest (14) were broader ‘big picture’ reviews. The most common topics were interventions (42%) and exposure/etiology (32%), with no reviews of diagnostic tests. The majority of the SRs had very low (71%) or low (19%) credibility, and few reached a high (7%) or moderate (3%) credibility. The most common issues were limitations with protocols, limited search strategies, and failure to account for risk of bias in drawn conclusions.

**Conclusions:** Few doctoral students included SRs in their theses, and the few SRs included in doctoral theses generally had a low credibility. Increasing the rate and quality of SRs in doctoral theses can help improve quality and relevance of subsequent primary research, and help students develop important skills. Actions are needed to support doctoral students to conduct high quality SRs.

**What is new?:**

- Few doctoral students included systematic reviews (SRs) in their theses
- The few SRs included in doctoral theses generally had a low credibility
- Increasing the rate of SRs can help improve the relevance of subsequent research
- Moreover, to support development of important skills and reach educational goals
- Actions are needed to support doctoral students to conduct high quality SRs

## Background

Systematic reviews are essential to ensure that decisions are informed by an up-to-date and complete understanding of the relevant research evidence (Higgins 2023). A systematic review attempts to find, assess and summarize all the empirical evidence that fits pre-specified eligibility criteria in order to answer a specific research question and can be conducted within a diverse set of areas that includes, but are not limited to, interventions, diagnostic tests, prognostic models and qualitative studies.

In 2022, approximately 6390 students were enrolled in a doctoral program in medicine and health at Swedish universities, accounting for 36% of all doctoral students across all research fields (Statistiska Centralbyrån 2024). The main aim of doctoral education is to “develop the knowledge and skills required to be able to undertake autonomous research” (Swedish Council for Higher Education 2024). Furthermore, specific goals must be reached within the education as determined by Swedish legislation, see Table 1. These substantially overlap with the learning process, application of tools and final output of a systematic review.

**Table 1:** Learning outcomes for doctoral students, stated in The Higher Education Ordinance, Annex 2.

|  |
| --- |
| **Table 1:** Learning outcomes for doctoral students, stated in The Higher Education Ordinance, Annex 2 |
| For the Degree of Doctor the third-cycle student shall:<br>• demonstrate **familiarity with research methodology** in general and the methods of the specific field of research in particular.<br>• **demonstrate the capacity for scholarly analysis and synthesis** as well as to **review and assess new and complex phenomena**, issues and situations autonomously and **critically**<br>• demonstrate the ability to identify and formulate issues with scholarly precision critically, autonomously and creatively, and to plan and use appropriate methods to undertake research and other qualified tasks within predetermined time frames and to **review and evaluate such work**<br>• demonstrate through a dissertation the ability to make a **significant contribution to the formation of knowledge** through his or her own research<br>• demonstrate the ability in both national and international contexts to present and discuss research and research findings authoritatively in speech and writing and in dialogue with the academic community and society in general<br>• demonstrate the ability to **identify the need for further knowledge** and<br>• demonstrate the capacity to contribute to social development and support the learning of others both through research and education and in some other qualified professional capacity.<br>• demonstrate intellectual autonomy and disciplinary rectitude as well as the ability to make **assessments of research ethics**, and<br>• demonstrate specialised insight into **the possibilities and limitations of research**, its role in society and the responsibility of the individual for how it is used. |

A summary of the potential benefits of conducting a systematic review is presented in Table 2. A systematic review can identify all relevant research to inform future projects, and transparently, comprehensively and unbiased summarize the current state of knowledge. The importance of conducting a systematic review before starting new research has previously been discussed extensively (Clarke 2010).

**Table 2:** Potential benefits of conducting a systematic review.

| Table 2: Potential benefits of conducting a systematic review |
| --- |
| • Identify all relevant research and thereby mitigating bias from selective outcome reporting and publication bias<br>• Systematic mapping and creating a summary of all previous knowledge within a specific topic<br>• Learn the strength and limitations of all previous studies within a specific topic<br>• Learn how to critically appraise and assess uncertainty in previous studies within a specific topic<br>• Find knowledge gaps where new research is needed – and thereby mitigating research waste and unnecessary duplication<br>• Generate additional research ideas based on previous studies and current evidence<br>• Identify ongoing studies, mitigating unnecessary duplication and inviting collaboration<br>• Avoid the use of previous research without rigorous critical appraisal and assessment of risk of bias |

For example, including a systematic review in a doctoral thesis can inform and improve the quality and relevance of subsequent studies (Glasziou 2023). A systematic review can also identify knowledge gaps and any ongoing studies on the same topic. This can help reduce duplication and limit redundant, harmful, and unethical research (Chalmers 2014) (Kim 2020) (Mahtani 2016b) (Clarke 2014). This, in turn, can limit research waste. Indeed, as much as 85% of biomedical research has been estimated to be research waste, where failure to build on current knowledge is a major contributor (Glasziou 2018).

Conducting a systematic review can also help doctoral students acquire important skills to critically consume and use research (Mahtani 2016a), identify and learn from the challenges and limitations of previous studies (Nikolakopoulou 2019), and learn to assess publication or reporting biases (Chalmers 2009). At the end of the doctoral period, a systematic review can put the generated knowledge into context by relating it to all other knowledge on the same topic. A systematic review can also facilitate the translation of the findings and be used as a basis for evidence-based decision making in healthcare, aiding in implementation or de-implementation, and facilitating the use of the best available evidence in clinical decision making.

The aim of this study was to map the extent of systematic reviews in doctoral theses at Swedish medical faculties, map their topic area, and assess the credibility of their findings.

## Objective

To explore the extent to which systematic reviews are included in doctoral theses from all medical faculties in Sweden, describe the type and topic and assess the credibility of the reviews.

## Methods

This cross-sectional study adhered to the STROBE reporting guidance. A prespecified protocol was published before searches, data extraction, or analyses began (Ringsten 2023). Search results, all data extracted, and analyses performed are available in an open repository (Ringsten 2023).

### Inclusion criteria

We included all doctoral theses published in 2021 at seven universities with a medical faculty in Sweden; Lund University; Uppsala University; Karolinska Institute; Gothenburg University; Linköping University; Umeå University; and Örebro University. Any reviews of the current literature using a systematic search and a structured research question were eligible, and the use of additional systematic methods was assessed in a separate step (see ‘Critical appraisal’-section). This included both systematic reviews, as defined in the Cochrane Handbook (Higgins 2023), and reviews within the broad ‘big picture review’-family (Campbell 2023).

### Search strategy

Search strategies were developed and run by information specialists within the university library at each university from May to September, 2023. The searches and databases were tailored for each medical faculty due to differences in local repositories. Doctoral theses were available in a full-text format for all universities. When a full-text thesis was not available in digital format, a physical version of the thesis was identified through archives. All the identified theses with links to repositories are available in the Supplementary material.

### Screening and data extraction

Screening and extraction forms were initially piloted for Lund University theses. Each thesis was screened for any systematic reviews independently and in duplicate in the title, abstract, the full list of conducted studies during the doctoral period, summary of the thesis content and, if needed, in full-text. Disagreements were resolved by consensus involving a third reviewer. Any uncertainties regarding the study design in title, abstract, study list and summary-stage of the thesis led to a full-text assessment.

All identified reviews within the theses were screened for any separate publication in a scientific journal following the references of the systematic review within the thesis. If no reference was available within the thesis to a scientific publication, the first author’s name was identified, and the publishing record of this author was further scrutinized for a scientific publication of the systematic review. More comprehensive reporting in journal articles was used, if available, in assessments of review type, topic, and credibility. Data extraction was done in duplicate by two independent reviewers. Any discrepancies between the two reviewers, and additional uncertainties, were resolved by discussion and consensus within the author team. Identified reviews included in doctoral theses were categorized based on “review type” (i.e. systematic reviews or broad reviews), and systematic reviews were additionally categorised based on the topic of the review. Furthermore, data was collected for how many studies were identified in the full search within the systematic reviews, number of included studies in the synthesis, software used for screening, tools used for critical appraisal and meta-analysis, and whether the GRADE framework was used.

### Critical appraisal

Each identified systematic review was assessed independently and in duplicate by two reviewers using the AMSTAR-2-tool to assess the methodological rigor and credibility of findings for each review (Shea 2017). For non-intervention systematic reviews where not all AMSTAR-2 items were applicable, the tool was adapted to fit these other topics based on previous work on the adaption of AMSTAR-2 (Puljak 2023), see Supplementary material for more information on how this was done. Any discrepancies between the reviewers were resolved by discussion, and consensus for judgements was reached within the author team.

### Outcomes

Our primary outcomes were the proportion of PhD theses, including at least one systematic review, and the proportion of systematic reviews of all articles included in the thesis. Our secondary outcomes were the type of review (systematic review and broad reviews), review category (e.g. intervention, exposure, prevalence), and the overall credibility of systematic review findings as assessed by AMSTAR-2. A number of prespecified exploratory outcomes were also analyzed, including: the tools used to support conduction of the systematic review and time to publication from the thesis publication date. Three post-hoc exploratory outcomes were also added during data extraction, including; the number of identified studies in the searches, the final number of included studies in the syntheses and the publication status of the reviews.

### Statistical analysis

Descriptive statistics was used to present the primary and secondary outcomes with numbers and percentages. In addition, the exploratory outcomes also included presentation of means with standard deviation and range.

## Results

Our search identified 852 theses published at Medical Faculties in Sweden in 2021, including 3461 studies in total. Of these studies, 45 were reviews of any type (1.3% of all studies), and 45 doctoral theses included a review (5.3% of all theses). A flow diagram is available in the supplementary material. The proportion of reviews varied among universities, from 3.4% of all studies at Örebro University to 0% of all studies at Linköping University (Table 3).

**Table 3:** Prevalence of reviews from medical faculties at Swedish universities.

|  |  |  |
| --- | --- | --- |
| **University** | **Proportion of studies that is a review** | **Proportion of theses with a review** |
| Overall across universities | 1.3% (45 reviews of 3461 studies) | 5.3% |
| Umeå University | 2.4% (6 reviews of 246 studies) | 9.8% |
| Karolinska Institute | 1.3% (18 reviews 1347 studies) | 5.3% |
| Lund University | 0.4% (3 reviews of 696 studies) | 1.8% |
| Göteborg University | 1.8% (10 reviews of 551 studies) | 7.3% |
| Uppsala University | 1.0% (4 reviews of 385 studies) | 4.3% |
| Örebro University | 3.4% (4 reviews of 116 studies) | 14.3% |
| Linköping University | 0.0% (0 reviews of 120 studies) | 0.0% |

Almost all (98%; 44 out of 45) of the identified reviews were published in a scientific journal. The median number of screened records was 979 (IQR 2346) and the median number of included studies was 13 (IQR 18).

Thirty-one (69%) of the reviews were systematic reviews according to our pre-defined criteria, and 14 (31%) were other types of reviews (e.g., broad “big picture” reviews). Out of the 31 systematic reviews, 13 (42%) explored the effects of interventions, 10 (32%) exposure and etiology, 3 (10%) qualitative research, 2 (7%) prevalence and incidence, 2 (7%) prognostic factors and prediction models, and 1 (3%) methodology.

The broad “big picture” reviews had very broad aims and inclusion criteria, and categorizing these into distinct topic areas was not possible (all these are listed in the Supplementary material).

### Credibility of the systematic reviews

Credibility of reviews were generally poor. Across all systematic reviews, 2 (7%) had high credibility and 1 (3%) had moderate credibility. In comparison, 6 (19%) had low credibility and 22 (71%) had critically low credibility with one or several critical weaknesses.

The most common methodological weaknesses contributing to lower credibility of the systematic reviews were issues with or lack of protocols (65%), limited search strategies (65%), and a failure to account for risk of bias in drawn conclusions (52%). All assessments and detailed justifications are reported in the Supplementary material.

### Tools used within the systematic reviews

Most (87%; 27 of 31) systematic reviews, conducted risk of bias assessments. The most common tools were the Cochrane Risk of bias-tools, i.e. RoB1 or RoB2 (used in 39% of systematic reviews), and Newcastle-Ottawa Quality Assessment Scales (NOQAS) (used in in 25% of systematic reviews). Five systematic reviews (15%) used the GRADE approach to assess the certainty of the evidence. The most common software used for meta-analysis was RevMan which was used in 7 systematic reviews (35%), while 5 (25%) used Stata, and 3 (15%) used R. Five (16%) of the systematic reviews reported the use of any specific software in the screening process (e.g., such as Covidence).

Additional results for proportion of published reviews, time to publication, use of software and specific tools in the review process are available in the Supplementary material.

## Discussion

We found that only 1.3% of all studies included in doctoral theses from medical faculties in Sweden were reviews of any type, and only 5.4% of all doctoral theses included a review. Most of these were of low methodological quality: only 10% of the systematic reviews had a high or moderate credibility. There is no consensus regarding the optimal proportion of reviews in theses. Nevertheless, we believe that our findings, in light of the many potential benefits of doctoral students performing a systematic review as part of their thesis (Table 2), warrants efforts to increase the number of systematic reviews included in doctoral theses. For example, a higher rate of systematic reviews in doctoral theses would help reach the learning outcomes of a doctoral education, avoid research waste, identify knowledge gaps, gain skills to assess bias and quality, and create a trustworthy synthesis of the research evidence within a particular field.

Not all doctoral theses benefit from including a systematic review. If there is already an available, up-to-date, relevant, comprehensive, high quality-systematic review within the specific topic of the thesis, it would be a waste of resources to duplicate this effort. Indeed, with an increase in the production of systematic reviews over the last few decades (Siontis 2018), overlapping reviews have been identified as research waste (Siontis 2018) (Page 2016) (Hoffman 2021). At the same time, systematic reviews of high quality are still not the norm (Page 2016) (Hoffman 2021) and the development of new methods for systematic reviews could lead to more accurate and nuanced results. Furthermore, as new evidence emerges, new or updated systematic reviews are needed to provide up-to-date evidence, and as the total number of available systematic reviews has increased, so have the available topics and types of reviews (Hoffman 2021).

Previous methodological research has shown that different methodological choices within reviews can have a large impact on results and conclusions (Sandau 2023). Replication of impactful research, such as systematic reviews, would improve, or refute the credibility of any conclusions drawn. Purposeful replication of systematic reviews is argued to play an important role in evaluating the credibility of previously generated knowledge (Karunananthan 2023) and guidance on when a systematic review should be replicated (Tugwell 2020), updated or adapted (Glasziou 2023) is available.

There are many potential barriers and facilitators to conducting systematic reviews within a doctoral thesis (McLennan 2021). Most notably are the policy, rules and culture for including reviews within a thesis. A summary of the barriers and facilitators that we have encountered in the process of performing this study is presented in table 4.

**Table 4:** Potential barriers and facilitators for conducting systematic reviews within doctoral theses.

| Table 4: Potential barriers and facilitators for conducting systematic reviews within doctoral theses |
| --- |
| • Available resources such as infrastructure, support and training. Potential activities to promote, improve or implement resources includes:<br>• Courses, workshops, seminars and training in conduction of high-quality systematic reviews and the methods that needs to be used<br>• Search strategy support by trained information specialists<br>• Methods support<br>• Summaries and guidance to resources for systematic reviews<br>• Infrastructure to support review production, e.g. Covidence for screening, RevMan Web for reporting and conducting meta-analyses, and Cochrane Interactive Learning for training.<br>• The policies, cultures and mindsets in medical faculties. Potential activities to promote, improve or implement ways to change system-related factors includes:<br>• A clear policy regarding the inclusion of systematic reviews in PhD theses.<br>• Dissemination of policies about the inclusion of systematic reviews in PhD theses<br>• Encouragement to conduct systematic reviews from supervisors, leaders of research groups and the medical faculty.<br>• Faculty-wide knowledge of what a systematic review is and what it can be used for. |

### The credibility of systematic review findings

The overall subpar methodological rigor in the systematic reviews indicates limitations in methodological understanding within the author teams, a lack of support from methodological expertise, or simply the lack of resources needed to conduct high quality reviews within Swedish universities. At the same time, the presence of a small number of systematic reviews rated as having high or moderate credibility, without any critical flaws, shows that it is possible to create high quality systematic reviews for doctoral students and include them in a thesis. The credibility of the findings of systematic reviews has been explored in several previous studies within other samples of systematic reviews, and they generally mirror the results of this study. The rating of very low credibility for systematic reviews has been estimated to be 74% in reviews of low-back pain (Almeida 2020), 88% within reviews of interventions after cancer (Siemens 2021), 88% for reviews of treatment after major depression (Matthias 2020), and 95% for reviews indexed in PsychINFO (Leclercq 2020).

### Identification of broad big picture reviews

A total of 31% of all reviews identified were broad big-picture reviews, which was a surprisingly high proportion. This might reflect the uncertainty of what characterizes a systematic review and vague policies for what constitutes a review that can be included in doctoral theses. The policies for inclusion of reviews in theses will be explored in an upcoming project and discussions within universities are needed to explore how policies can define how and if broad big-picture reviews can be included in doctoral theses.

### Strengths and limitations of this study

We are not aware of any other studies exploring the extent of reviews included in doctoral theses. How Sweden compares to other countries needs to be explored further.

The main strengths of this study include the inclusion of all medical faculties within Sweden and involving collaborators from these faculties. Furthermore, the project was co-designed, co-executed, and facilitated by doctoral students, ensuring an end-user perspective of the process and output. In addition, duplicate assessments and extractions, use of a pre-registered protocol, sharing of data to facilitate reuse, and transparency enhance robustness and transparency.

A limitation of our study is that we did not evaluate the need for a systematic review (i.e., if any systematic review was conducted outside of the doctoral theses, or if credible systematic reviews by other researchers was used as a basis for subsequent research). Nevertheless, we believe that the very low rates of systematic reviews in doctoral theses identified in our study warrants efforts to increase this number even in the absence of such judgements.

The application of AMSTAR-2 to interventions includes interpretation and judgment (DeSantis 2023) (Gates 2020) which is an inherent limitation of any critical appraisal. Within this study 6 out of 45 (13%) final credibility judgements differed between the two reviewers needing to be solved by discussion and consensus, which is considered to be in the higher spectrum of inter-rater reliability (Shea 2017).

Furthermore, the modification of AMSTAR-2 to non-intervention reviews, which it is not originally designed for, could have an impact on the final judgements – but would bias our results towards higher credibility and could therefore our results should be viewed as a conservative assessment. Extensions to AMSTAR-2 for other types of studies are currently being planned (Puljak 2023), and the evaluation of critical appraisal tools for broad review types is ongoing (Pollock 2022) and could be used in updates or within similar projects.

## Conclusions

Few doctoral students conduct and include systematic reviews in their theses, and hence do not get experience in using systematic review-methods or to systematically evaluate their field of study. The low credibility of the conducted SRs is also a concern.

Increasing the rate and quality of SRs in doctoral theses can help improve quality and relevance of subsequent primary research, and help students develop important skills. If conducting high quality SRs are considered an important goal in the training of the future generation of researchers - actions are needed to support doctoral students to conduct high quality SRs.

## Supporting information

Supplementary material 1: Search strategy, flowchart, modified AMSTAR-2, additional results, deviations from intended methods

Supplementary file 2: Review characteristics, AMSTAR-2 assessments, list of all searched theses

## Data Availability

All data produced are available online at Open Science Framework doi.org/10.17605/OSF.IO/DRJ6Q

## Acknowledgements

We would like to thank the project group, including representatives from all Medical Faculties in Sweden in the support with developing search strategies in local repositories and support in screening of doctoral theses, including: Lea Styrmisdottir, Sabina Gillsund, Narcisa Hannerz, Emma-Lotta Säätelä, Linda Hammarbäck, Katarina Jonzon, Isolina Ek, Karolina Schröder, Jenny Aspling Rydgren, Jenny Betmark, Joakim Westerlund, Görel Sundström, Malin Barkelind, Matilda Naesström, Viktoria Johansson, Mattias Lennartsson, Ann Emilsson, Camilla Gothberg and Liz Holmgren.

We would also like to greatly thank Maria Björklund for feedback on the protocol and for support in setting up contacts with libraries at all Faculty of Medicine in Sweden.

## Declaration and conflicts of interests

MR affiliated to Cochrane Sweden, and currently receive salary for creating and teaching related to systematic reviews in university settings and within Swedish government organizations.

MJ is affiliated to Cochrane Sweden and declare no other conflict of interest.

MB affiliated to Cochrane Sweden, and currently receive salary for creating and teaching related to systematic reviews in university settings and within Swedish government organizations.

KF Declares no conflict of interest.

## CRediT Author statement

Martin Ringsten: Conceptualization, Formal analysis, Methodology, Data Curation, Writing - Original Draft, Project administration

Minna Johansson: Conceptualization, Formal analysis, Methodology, Writing - Original Draft

Matteo Bruschettini: Conceptualization, Formal analysis, Methodology, Writing - Review & Editing

Kenneth Färnqvist: Formal analysis, Methodology, Writing - Review & Editing

