## Supplementary material 1: Search strategy, flowchart, modified AMSTAR-2, additional results, deviations from intended methods for "Inclusion, characteristics and credibility of systematic reviews in doctoral theses: A cross-sectional study of all Medical Faculties in Sweden"

**Supplementary material file 1**

**Appendix 1: Search strategies**

- Umeå University was searched through the DiVA-portal (<http://umu.diva-portal.org>)
- Karolinska University was searched through KI Open Archive (<https://openarchive.ki.se>)
- Lund University was searched through Lund Research Portal (<https://portal.research.lu.se/en/publications>)
- Göteborg University was searched through Gothenburg University Publications Electronic Archive (<https://gupea.ub.gu.se>)
- Uppsala University was searched through the DiVa-portal (<https://uu.diva-portal.org>)
- Örebro University was searched through the DiVa-portal (<https://oru.diva-portal.org>)
- Linköping University was searched through the DiVa-portal (<https://liu.diva-portal.org>)

All repositories were filtered for theses (all sorts) published in 2021 at the Faculty of Medicine.

All screened theses and the identified theses with a review for each university is openly available in the repository at <https://osf.io/drj6q/>.

**Appendix 2: Flowchart**

**Identification of theses via databases and registers**

Records removed *before screening*:

Duplicate records removed (n = 0)

Records marked as ineligible by automation tools (n = 0)

Records removed for other reasons (n = 0)

Studies identified from databases and registries in 2021 at:

Lund University (n = 696)

Karolinska Institute (n = 1347)

Umeå University (n = 246)

Göteborg University (n = 551)

Uppsala University (n = 385)

Örebro University (n = 116)

Linköping University (120)

**Identification**

Studies excluded:

No review methods used (n = 3416)

Studies screened

(n = 3461)

**Screening**

Theses with an included review

(n = 45)

Studies with review methodology (n = 45 )

**Included**

**Appendix 3: Applying and modification of AMSTAR-2**

AMSTAR-2 was chosen based on the time to complete an assessment, reported higher agreement between reviewers (Gates 2020) and review team's familiarity of the tool.In short, three questions deemed not applicable to non-intervention reviews were removed (item 1, item 3 and item 8) and four questions were revised to only signal the inclusion of a certain methodological step and not assessing exactly how it was carried out (item 2, item 9, item 11 and item 15). The adaptions were made in order to more accurately compare and summarise methodological choices and credibility between systematic reviews conducted in different topics using the same base for appraisal, and to rely on previous work using a commonly used instrument.

Item 2. Did the report of the review contain an explicit statement that the review methods were established prior to the conduct of the review and did the report justify any significant deviations from the protocol? (Yes, Partial Yes, No). Our guidance: just state (yes/no) if a protocol was established or not, and not go into the actual content that the guidance explains in more detail.

Item 9. Did the review authors use a satisfactory technique for assessing the risk of bias (RoB) in individual studies that were included in the review? (Yes, Partial Yes, No). Our guidance: Just state (yes/no) if RoB was assessed or not, and not detailed what domains was considered.

Item 11. If meta-analysis was performed did the review authors use appropriate methods for statistical combination of results? (Yes, No, No Meta-analysis Conducted). Our guidance: Just state (yes/no) if meta-analysis was used a model for synthesis or not.

Item 15. If they performed quantitative synthesis did the review authors carry out an adequate investigation of publication bias (small study bias) and discuss its likely impact on the results of the review? (Yes, No, No Meta-analysis Conducted). Our guidance: Just state if publication bias was considered in any way in the report.

**Appendix 4: Specific additional results**

44 out of 45 (98%) of all review types were published in a scientific journal. 36 (81%) were published in a scientific journal before the publication of the thesis. Of the 8 (18%) reviews published after publication of the thesis, the median time was 263 days (IQR 324). The number of screened records was median 979 (IQR 2346) with a minimum of 69 and maximum of 13606 within the systematic reviews. The final number of included studies was median 13 (IQR 18), minimum 3, maximum 215.

Only 5 (16%) systematic reviews reported the use of specific software in the screening process. 2 (7%), used Endnote and 1 (3%) used Covidence, 1 (3%) used Rayyan and 1 (3%) used Excel. 20 systematic reviews (65%) used meta-analysis as a synthesis-method, and 11 (35%) used a mix of tabular and narrative formats for synthesis. The software used for meta-analysis was 7 (35%) RevMan, 5 (25%) used Stata, 3 (15%) used R. Only one systematic review used either SAS (5%), Comprehensive Meta-analysis package (5%), or StatsDirect 3 (5%), while two reviews did not specify the software used.

27 out of 31 (87%) systematic reviews conducted risk of bias assessments, and only one used tools from different assessment-families. Out of the systematic reviews, 11 (39% %) used the Cochrane Risk of bias-tools (RoB1 or RoB2), 7 (25%) used Newcastle-Ottawa Quality Assessment Scales (NOQAS), and 2 (7%) used Critical Appraisal Skills Programme Qualitative Studies Checklist (CASP). Only one systematic review used either QUADRAS-2, Scottish Intercollegiate Guidelines checklist, MINORS, the SYRCLE RoB tool, Joanna Briggs Institute (JBI) Critical Appraisal tools, Downs and Black checklist, Drummond Checklist for Economic Evaluation of Health Care Programmes, or the SBU assessment tool for qualitative studies. Only 5 systematic reviews (15%) used the GRADE approach to assess certainty of evidence.

**Appendix 5: Deviations and updates from intended methods:**

After piloting the initial data extraction from the identified reviews, we saw the need to create distinct review type categories in the “Other” category. This includes reviews of; Risk/Etiology and Methods/Models.

We did not pre-specify to report the extent reviews that were published in a scientific journal and the timing of publication, but noticed during piloting-process of the data extraction that we were able to easily assess and calculate these numbers. We believe that a post-hoc decision to include these numbers would give additional nuance to the study, and have included these numbers in the result section.

The larger than expected proportion of “non-systematic reviews” (e.g. scoping reviews and reviews with broad research questions) coupled with the lack of consensus regarding categorisation of these reviews prompted us to define these reviews being within a “Broad review family”.

The outcome of presenting the results from the credibility of findings-assessment with AMSTAR-2 was not prespecified in the protocol in the outcome domain, but was specified in as an objective and the methods. For clarify we added this as a secondary aim to the study.
